# Design differences explain variation in results between randomized trials and their non-randomized emulations

**DOI:** 10.1101/2023.07.13.23292601

**Authors:** Rachel Heyard, Leonhard Held, Sebastian Schneeweiss, Shirley V Wang

**Affiliations:** Center for Reproducible Science, Epidemiology, Biostatistics and Prevention Institute, University of Zurich, Hirschengraben 84, 8001 Zurich, Switzerland; Division of Pharmacoepidemiology and Pharmacoeconomics, Department of Medicine, Brigham and Women’s Hospital, Harvard Medical School, 1620 Tremon St, Boston MA 02120

**Author notes:** **Corresponding Author:** Rachel Heyard, PhD, EBPI, Hirschengraben 84, 8001 Zürich, Switzerland.

**Keywords:** Real-world evidence, randomized controlled trial, design, emulation differences, meta-regression, heterogeneity

## Abstract

**Objectives:** While randomized controlled trials (RCTs) are considered a standard for evidence on the efficacy of medical treatments, non-randomized real-world evidence (RWE) studies using data from health insurance claims or electronic health records can provide important complementary evidence. The use of RWE to inform decision-making has been questioned because of concerns regarding confounding in non-randomized studies and the use of secondary data. RCT-DUPLICATE was a demonstration project that emulated the design of 32 RCTs with non-randomized RWE studies. We sought to explore how emulation differences relate to variation in results between the RCT-RWE study pairs.

**Methods:** We include all RCT-RWE study pairs from RCT-DUPLICATE where the measure of effect was a hazard ratio and use exploratory meta-regression methods to explain differences and variation in the effect sizes between the results from the RCT and the RWE study. The considered explanatory variables are related to design and population differences.

**Results:** Most of the observed variation in effect estimates between RCT-RWE study pairs in this sample could be explained by three emulation differences in the meta-regression model: (i) in-hospital start of treatment (not observed in claims data), (ii) discontinuation of certain baseline therapies at randomization (not part of clinical practice), (iii) delayed onset of drug effects (missed by short medication persistence in clinical practice).

**Conclusions:** This analysis suggests that a substantial proportion of the observed variation between results from RCTs and RWE studies can be attributed to design emulation differences.

**What is already known on this topic:** Real-world evidence (RWE) studies can complement randomized controlled trials (RCT) by providing insights on the effectiveness of a medical treatment in clinical practice. Concerns about confounding have limited the use of RWE studies in clinical practice and policy decisions.

**What this study adds:** A large share of the observed variation in results between RCT-RWE study pairs could be explained by design emulation differences.

## 1 INTRODUCTION

Real-world evidence (RWE) has been defined as evidence on the effects of medical products that are derived from the analysis of real-world data (RWD) which includes a variety of patient health data sources, particularly data collected as part of routine clinical practice, including electronic health records and insurance claims data [1]. There has been increasing interest in the use of RWE from RWD to support clinical practice and policy decisions [2–5]. However, there remain concerns about the validity of such evidence compared to the traditional randomized controlled trial (RCT) [5–7].

Such concerns stem from a misleading dichotomy that pits RCTs against database studies instead of viewing them as providing complementary information that informs a fuller understanding of drug effects [8]. There have been many efforts to compare database and RCT results. Some have observed high concordance and touted the ability of well-designed database studies to generate valid causal conclusions [9–13]. Others have used observed divergence in results to criticize database studies as intractably confounded [7,14–17].

One effort to compare RCT and database studies was RCT-DUPLICATE [10,19–21]. RCT-DUPLICATE set out to emulate more than 30 trials by prospectively designing a series of database studies to match each RCT design as closely as possible within the confines and limitations of using data that were not collected for research purposes. Because of the nature of using routinely collected data from clinical practice, some elements of trial design could not be exactly emulated, for example, measures to ensure prolonged adherence over long follow up windows. Such emulation differences can be summarised as: 1) differences in outcome measurements, 2) differences in demographics of included patients, 3) differences in treatment implementation in clinical practice, 4) lack of placebo in clinical practice. Design emulation differences change the question or estimand being addressed in the RCT compared to the database study [22,23].

We sought using the RCT-DUPLICATE collection of emulated trials to assess how design emulation differences relate to variation in results between RCTs and RWE database studies that were designed to emulate them. We explore whether the characteristics of emulation differences can reduce the residual heterogeneity in effect size differences in a meta-regression analysis.

## 2 DATA AND METHODS

The following analysis is of an exploratory nature and attempts to better understand emulation differences and how this impacts variation in results between RCT-RWE study pairs.

### RCT DUPLICATE

The selection process for the RCT-DUPLICATE study is described in detail elsewhere [19,24]. In summary, the RCT-DUPLICATE consortium emulated 32 RCTs that were relevant to regulatory decision making and were potentially feasible to emulate using claims data because key study parameters such as the primary outcome, the treatment strategies and inclusion/exclusion criteria were measurable. The selected trials include a mix of superiority and non-inferiority trials, trials with large and small effect sizes, and a mix of trials with active comparators and placebo added to active standard of care therapies. The consortium used three RWD sources to implement the database studies that emulated RCTs: Optum Clinformatics Data Mart beginning in 2004, IBM MarketScan beginning in 2003 and Medicare Parts A, B, and D. Whenever possible, the RCT emulations were implemented in more than one of the data sources and the final analyses are based on estimates resulting from a fixed effects meta-analysis of the implementations in all databases. In the present study only those trials where the primary analysis resulted in a hazard ratio will be used - the trial LEAD2 with continuous outcome is discarded. For two trials (ISAR-REACT5 and VERO) a Chi-squared test indicated that the results were heterogeneous across databases so that the meta-analysis could not be performed to obtain a pooled RWE estimate for the hazard ratio [19] and those trials are omitted too.

### Emulation differences identified in RCT-DUPLICATE

Design emulation differences were recorded as covariates in RCT-DUPLICATE. Differences in age and sex distribution are captured as numerical variables representing the difference in mean age or percentage females - the value in the RCT minus the value in the RWE pooled emulation. The categorical emulation difference characteristics recorded in RCT-DUPLICATE are described in Table 2.1 [19].

**Table 2.1:**
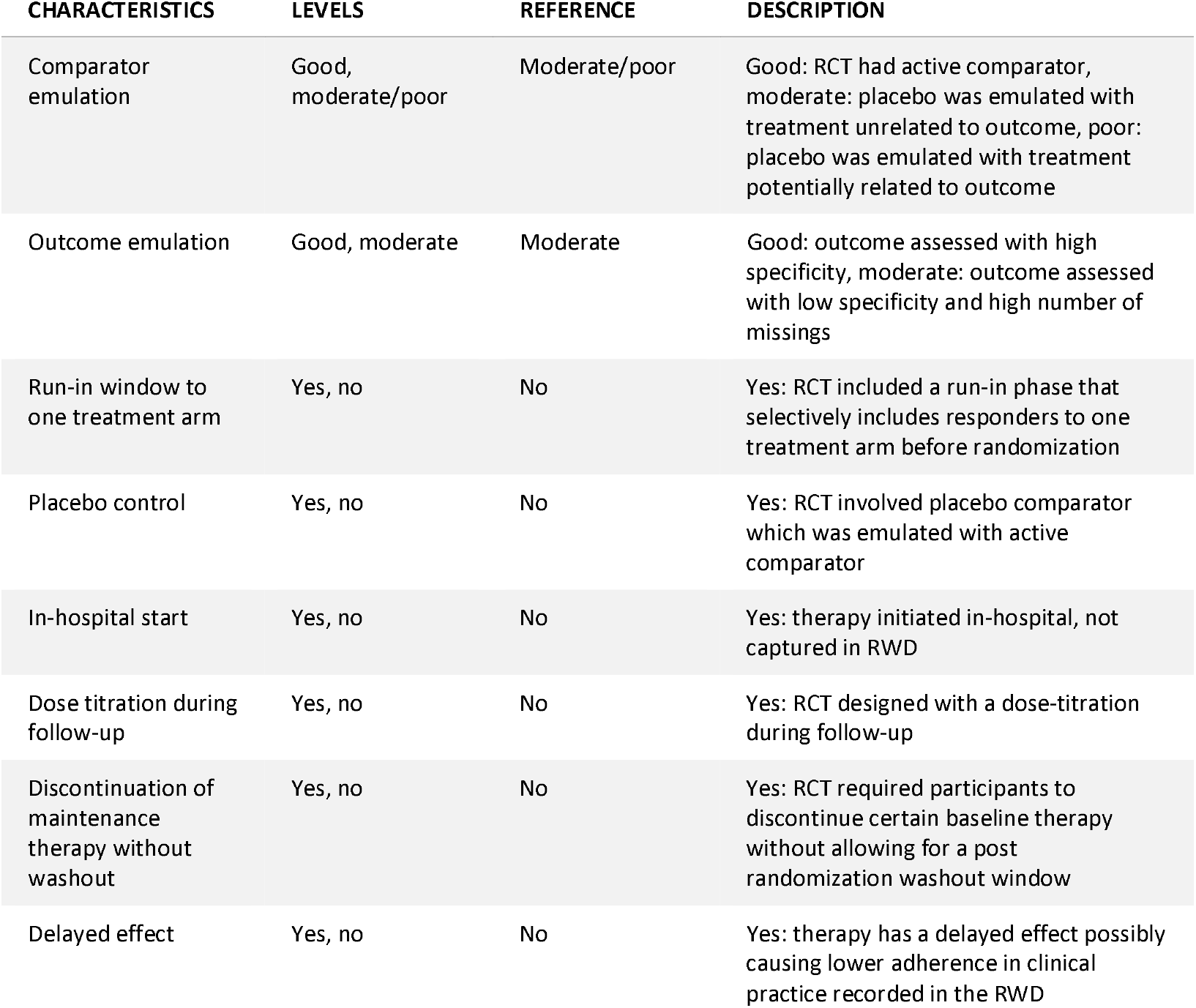
Description of categorical emulation difference characteristics with their possible levels, the reference category, as well as a short description are provided. All of the listed characteristics are binary.

| CHARACTERISTICS | LEVELS | REFERENCE | DESCRIPTION |
| --- | --- | --- | --- |
| Comparator emulation | Good, moderate/poor | Moderate/poor | Good: RCT had active comparator, moderate: placebo was emulated with treatment unrelated to outcome, poor: placebo was emulated with treatment potentially related to outcome |
| Outcome emulation | Good, moderate | Moderate | Good: outcome assessed with high specificity, moderate: outcome assessed with low specificity and high number of missings |
| Run-in window to one treatment arm | Yes, no | No | Yes: RCT included a run-in phase that selectively includes responders to one treatment arm before randomization |
| Placebo control | Yes, no | No | Yes: RCT involved placebo comparator which was emulated with active comparator |
| In-hospital start | Yes, no | No | Yes: therapy initiated in-hospital, not captured in RWD |
| Dose titration during follow-up | Yes, no | No | Yes: RCT designed with a dose-titration during follow-up |
| Discontinuation of maintenance therapy without washout | Yes, no | No | Yes: RCT required participants to discontinue certain baseline therapy without allowing for a post randomization washout window |
| Delayed effect | Yes, no | No | Yes: therapy has a delayed effect possibly causing lower adherence in clinical practice recorded in the RWD |

Finally, all characteristics from Table 2.1 are summarized into a binary composite covariate indicating whether the RCT-RWE study pair represents a ‘close emulation’ or not. A study pair is considered a ‘close emulation’ if the comparator and outcome emulations were at least moderate with at least one good, and none of the following: start of follow-up in hospital, a run-in window that selectively includes responders to one treatment arm, mixing effects of randomization and discontinuation of baseline therapy, and delayed effect over long follow-up. The composite indicator was defined retrospectively by the RCT-DUPLICATE team [19].

### Statistical analysis

All statistical analyses used require that the effect estimates from the RCT and the RWE are approximately normally distributed. Hence log-transformations are applied on the hazard ratios of all included studies. The standardized differences of the RCT-RWE study pairs are computed by dividing the difference in log hazard ratios by the standard error of the difference. The squared standardized difference is the *Q*-statistic which is used to perform the *Q* -test for heterogeneity between RCT and RWE studies [25,26]. The sum of all computed *Q* -statistics is used for an overall test for heterogeneity between RCT-RWE study pairs included in RCT-DUPLICATE.

Heterogeneity can be quantified as a multiplicative parameter [27], an overdispersion parameter generally larger than 1 inflating the model’s standard errors. As described in [28], the multiplicative heterogeneity parameter is estimated by fitting a weighted linear regression on the observed differences from all RCT-RWE study pairs against a constant, with weights defined as the inverse of the squared standard error of the differences. The multiplicative heterogeneity is then simply this model’s standard error and absence of heterogeneity is achieved if the parameter is equal to 1. Whenever the heterogeneity parameter is estimated to be smaller than 1, it is set to its lower bound of 1. Characteristics describing emulation differences are used to explain part of the observed heterogeneity. Using meta-regression methods ([29], Chapter 7), the emulation difference characteristics - the differences in age and sex distribution as well as the categorical characteristics summarized in Table 2.1 - are added into the weighted linear regression model estimating the multiplicative heterogeneity. If the extracted residual heterogeneity from the more complex, adjusted, model is smaller than the heterogeneity measured with the simple model (with only a constant), part of the variation can be explained by the included emulation difference characteristics. To reduce the complexity of the meta-regression, avoid overfitting and choose only the most predictive of the *p* candidate characteristics, leave-one-out cross-validated mean squared errors (LOO MSE, [30]) are computed for all 2^*p*^ possible candidate models. The simplest model with an MSE at most one standard error away from the smallest MSE across all models is selected [31]. The model coefficients for the included characteristics have to be interpreted with respect to the model’s intercept, the difference in RCT-RWE effect estimates that remains when all binary emulation difference characteristics and the centered continuous characteristics are set to their reference or zero, respectively.

A detailed description of the statistical analyses can be found in the appendix. All analyses are performed in the R programming language 4.3.0 [32]. Code and data to reproduce the analyses and recompile this manuscript are available through https://gitlab.com/heyardr/hte-in-rwe.

## 3 RESULTS

Figure 3.1 shows the estimated hazard ratios from the RCTs against the hazard ratios estimated using the pooled RWE studies with their 95% confidence intervals. Perfectly emulated trial estimates would scatter around the diagonal line. While more than half of the pooled estimates from the RWE studies tend to be smaller than the RCT estimates, many are also larger. This is different from the results seen in the large-scale replication projects where the effect size estimated in the replication study is generally smaller than the one from the original study, which may be attributable to publication bias or other questionable research practices which are unlikely operating in this emulation study [33]. This phenomenon is referred to as “shrinkage” of effect sizes [34], whereas we observe exaggeration in many of the emulations.

**Figure 3.1:**
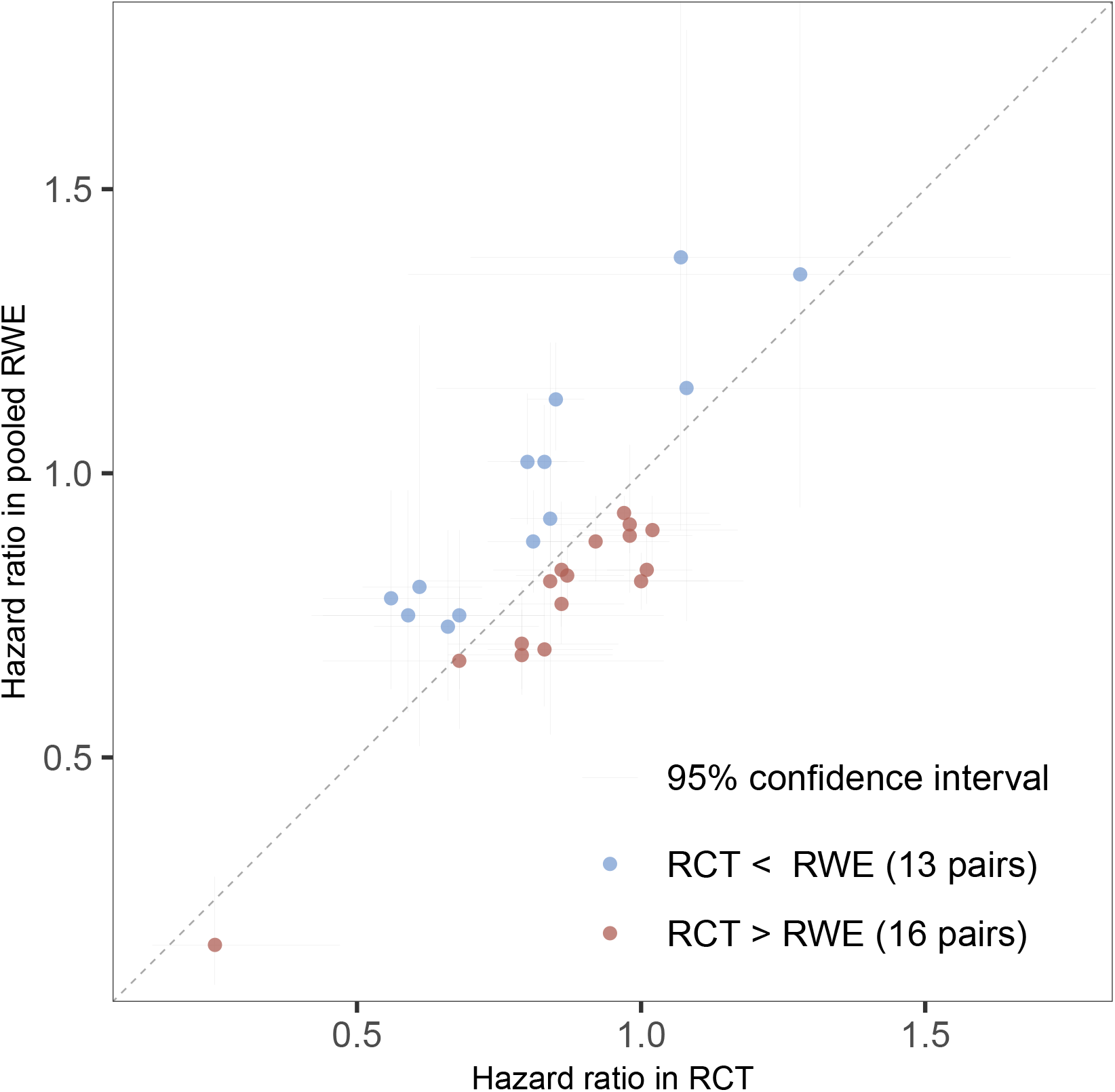
The hazard ratios estimated in the randomized trial and the RWE study (pooled over all data sources), together with their respective 95% confidence interval. The diagonal line represents a scenario of perfect emulation, while all trials with points on the right side of the diagonal have an effect size estimated in the RCT that is larger than the effect size estimated in the pooled RWE study.

The left panel of Figure 3.2 represents the distribution of the observed standardized differences versus a standard normal distribution, which should, in the absence of heterogeneity between each RCT and its emulation, be aligned. Then, the right panel of Figure 3.2 shows the *p*-values resulting from the *Q*-test for heterogeneity between each individual RCT-RWE study pair which would, in the absence of heterogeneity, be uniformly distributed. However, many small *p*-values are observed. The *p*-value from an overall test of heterogeneity suggests strong evidence for variation between all study pairs in RCT-DUPLICATE (*p* < 0.001).

**Figure 3.2:**
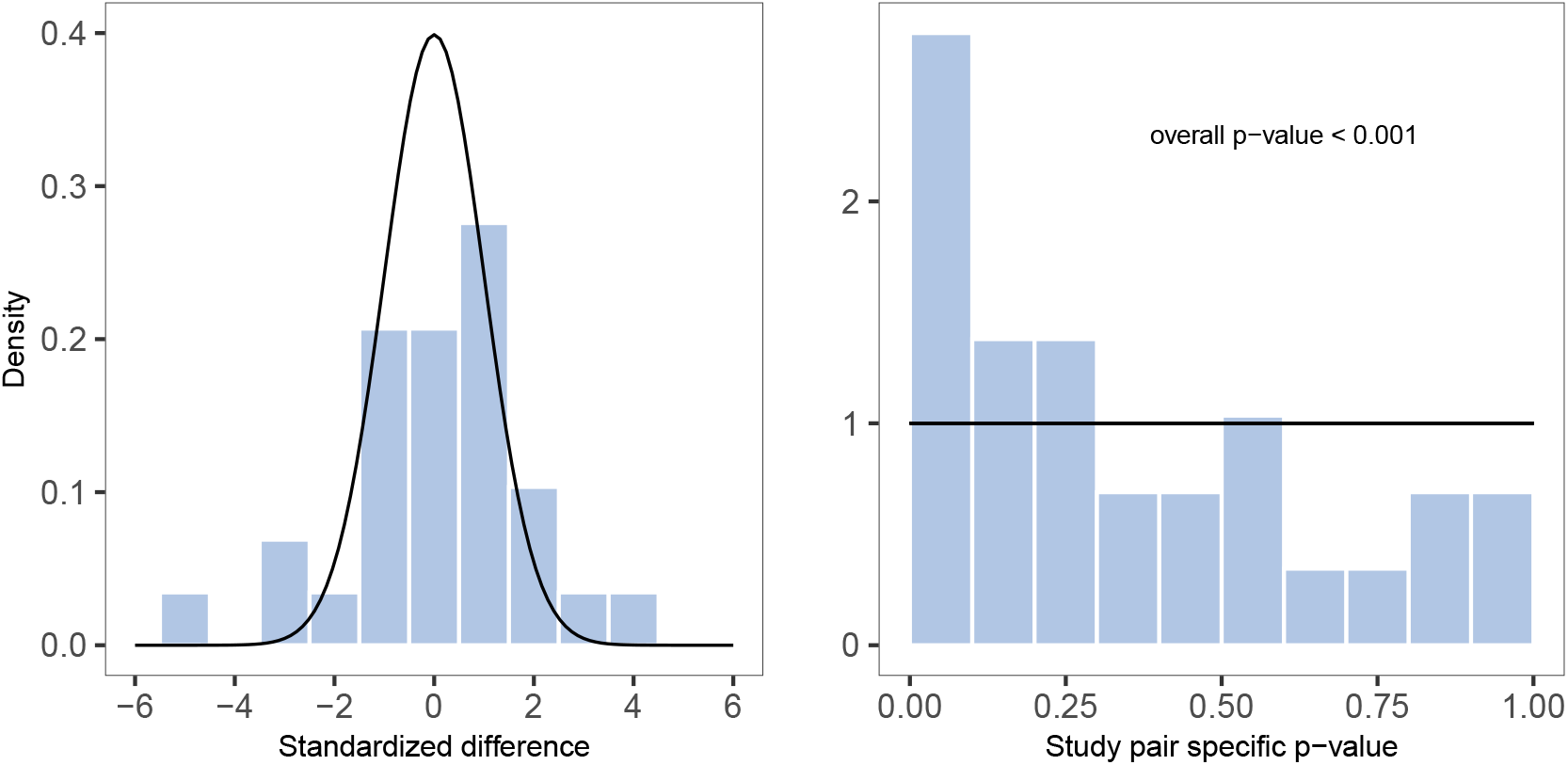
Left: The distribution of the observed standardized difference of the RCT-RWE study pairs compared to the standard normal distribution. Right: The p-values from the Q-test for heterogeneity within RCT-RWE study pairs compared to the uniform distribution on the interval [0-1], as well as the p-value for the overall test for heterogeneity between all study pairs.

To better understand the variability in results observed in RCT-DUPLICATE, the variation was quantified and its sources were investigated. Figure 3.3 represents the differences in log hazard ratio for each study pair depending on whether the emulation was categorized as close or not. Trials that are categorized as not closely emulated using the exploratory indicator tended towards positive differences. The estimated multiplicative heterogeneity comparing the pooled RWE studies to the RCT is shown in Table 3.1, together with the model intercept and coefficient with 95% confidence intervals. The simple model refers to the weighted regression with only a constant while the second model is a meta-regression adjusted for the binary characteristic “close emulation”. Including “close emulation” in the weighted linear regression model reduced the heterogeneity from 1.9 to 1.72. In other words, a small part of the observed variation between estimates in RWE-RCT study pairs can be attributed to the composite covariate. While the intercept of the simple model is close to zero, the intercept of the adjusted model - the difference in log hazard ratio for the trials that are not closely emulated - tends to be positive. Closely emulated trials however produce, on average, slightly negative differences. The same information can be found in the bubble plot in Figure 3.5.A.

**Table 3.1:** Model intercept and coefficient with their 95% confidence interval together with the heterogeneity between the RWE and the RCT depending on the model used. A heterogeneity close to 1 means more homogeneous effect size differences between study pairs.

| MODEL | INTERCEPT (95% CI) | COEFFICIENT (95% CI) | HETEROGENEITY |
| --- | --- | --- | --- |
| Simple | 0.002 (-0.06 to 0.07) | - | 1.90 |
| Adjusted for close emulation | 0.06 (-0.01 to 0.1) | -0.2 (-0.3 to -0.04) | 1.72 |

**Figure 3.3:**
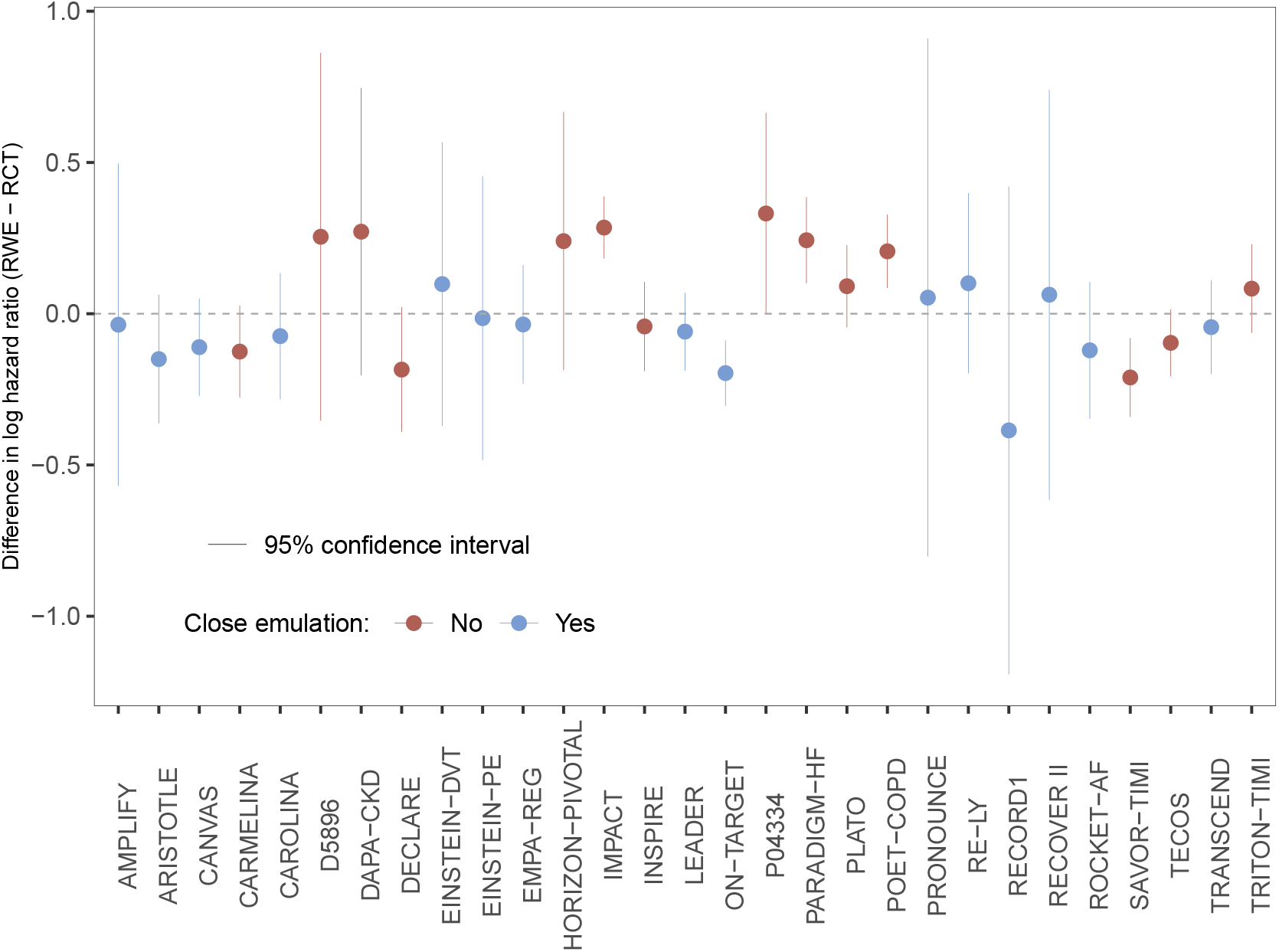
The difference in effect estimates, hazard ratios, observed in the RCT and the pooled RWE depending on whether it was a close emulation or not. The horizontal line represents a scenario of no difference between RWE and RCT estimates.

**Figure 3.4:**
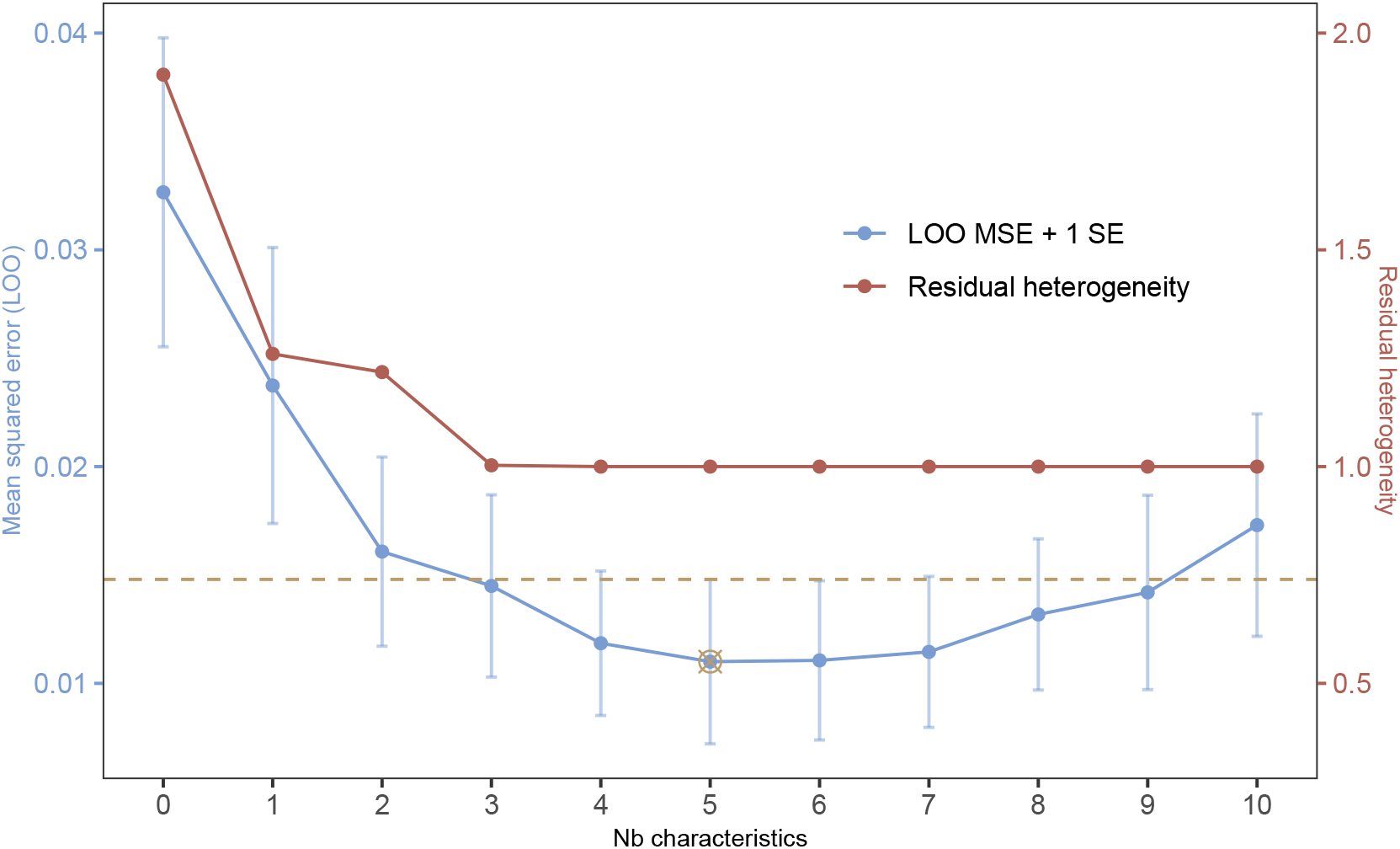
The leave-one-out mean squared error and the residual heterogeneity for each number of covariates included in the weighted regression. A multiplicative heterogeneity of 1 represents absence of heterogeneity in the effect size difference between study pairs. The minimum in MSE (best predictive performance) is found after including five characteristics. The simplest model with LOO MSE smaller than the minimum plus 1 standard error is the model with three characteristics.

**Figure 3.5:**
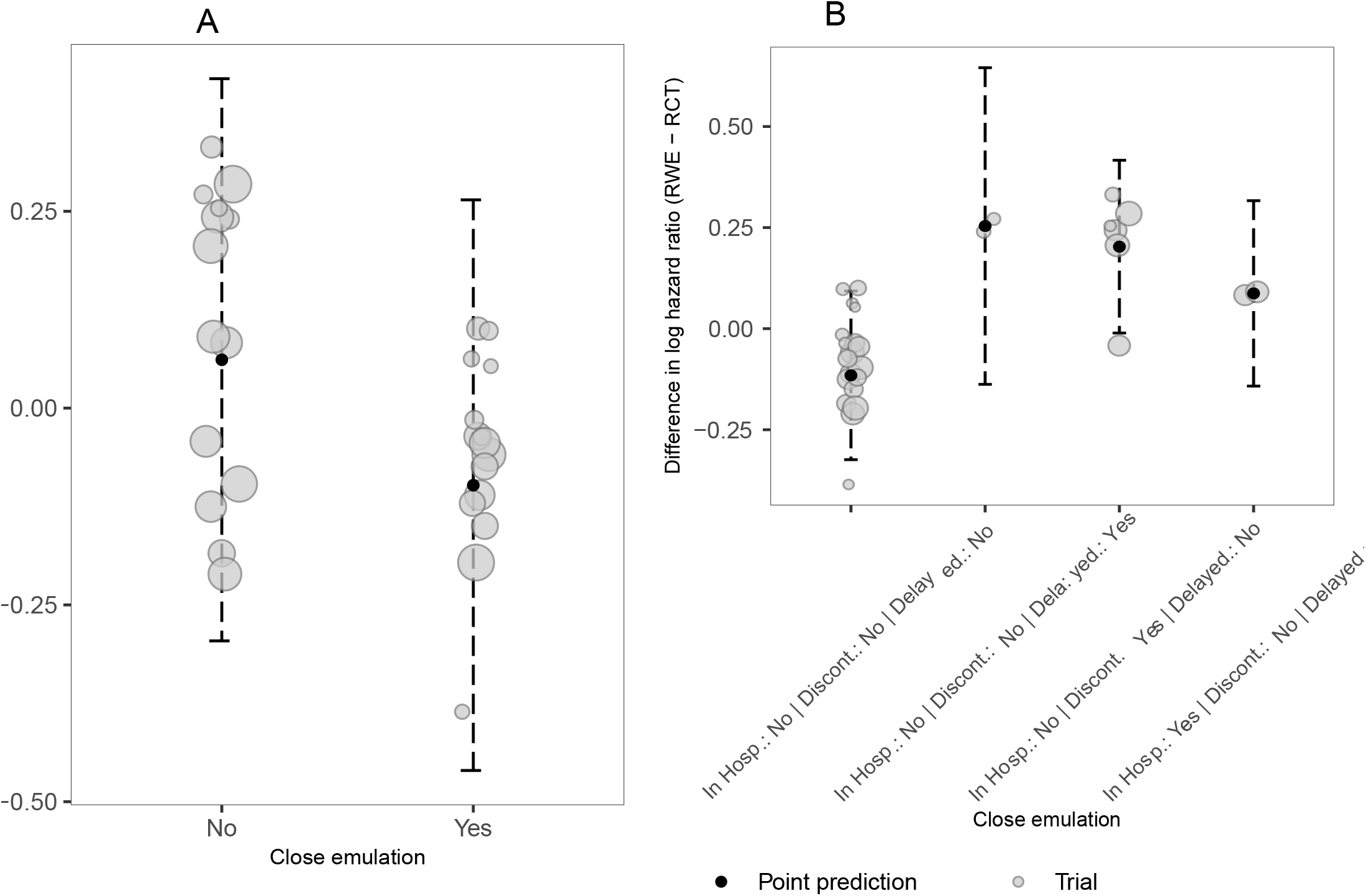
Bubble plots showing the association between close emulation (Yes or No) and the difference in log hazard ratio (A) and the association between the (possible) combination of the three binary characteristics and the difference in log hazard ratio (B). The larger the bubbles, the more precise the estimates/trials. Horizontal jitter has been applied on the bubbles to enhance visibility. The 95% prediction intervals are compiled using the meta-regression including the binary composite covariate.

A set of explanatory characteristics explaining emulation differences between RCT and emulation were explored to use instead of the composite covariate “close emulation”. Univariate model coefficients, the respective model intercept and the residual heterogeneity can be found in Table 3.2. Some of the characteristics reduced the heterogeneity more than others. Adding the characteristic “Discontinuation of maintenance therapy without washout”, for example, results in the largest decrease in heterogeneity: from 1.9 to 1.26. The intercept shown in Table 3.2 can be interpreted as the difference in log hazard ratio for the respective reference category of the binary characteristics or no difference in the distribution for the two continuous characteristics, age and percentage female. The variable selection algorithm described in the methods section was applied to decide which combination of the ten characteristics related to design emulation and population differences explains the most variation in the meta-regression. All 2^10^ = 1’024 possible candidate models, depending on which of the ten characteristics are included, were fitted and their LOO MSE were computed. The evolution of the LOO MSE depending on the number of characteristics in the model is illustrated in Figure 3.4. The residual heterogeneity from the respective models is shown. Note that for each number of characteristics, from 0 to 10, only the results from the model minimizing the LOO MSE are shown. A minimum is found after including five characteristics, with residual multiplicative heterogeneity of 1. Conditional on those five characteristics, differences in effect estimates are homogeneous. With a small sample size of only 29 included RCT-RWE study pairs, a reduction in the complexity of the meta-regression is desired in order to avoid overfitting. The final model is the simplest model with LOO MSE smaller than the minimum MSE observed in Figure 3.4 plus 1 standard error. Using this tuning parameter, three characteristics would be included. Table 3.3 shows the coefficient estimates of those models with best performance for each number of included characteristics. Note that the models summarised in Table 3.3 lead to the model performance and heterogeneity illustrated in Figure 3.4.

**Table 3.2:** Univariate coefficients estimate with 95% confidence interval for each candidate characteristics. For each row - each characteristics - a separate model is fit, resulting in a separate intercept and residual heterogeneity. The closer the residual heterogeneity is to 1, the more the characteristic explains part of the variations.

| CHARACTERISTICS | INTERCEPT (95% CI) | UNIVARIATE COEFFICIENT ESTIMATE (95% CI) | RESIDUAL HETEROGENEITY |
| --- | --- | --- | --- |
| Comparator emulation (good) | -0.06 (-0.1 to 0.03) | 0.1 (-0.005 to 0.2) | 1.82 |
| Outcome emulation (good) | -0.003 (-0.1 to 0.1) | 0.007 (-0.1 to 0.2) | 1.94 |
| Difference mean age (centered) | 0.009 (-0.06 to 0.07) | -0.01 (-0.03 to 0.01) | 1.91 |
| Difference in % female (centered) | 0.005 (-0.07 to 0.08) | 0.0007 (-0.005 to 0.007) | 1.94 |
| Run-in window to one treatment arm (yes) | -0.06 (-0.1 to 0.002) | 0.2 (0.1 to 0.4) | 1.51 |
| Placebo control (yes) | 0.07 (0.0001 to 0.1) | -0.2 (-0.3 to -0.06) | 1.68 |
| In-hospital start (yes) | -0.008 (-0.08 to 0.06) | 0.1 (-0.1 to 0.3) | 1.91 |
| Dose titration during follow-up (yes) | 0.02 (-0.05 to 0.1) | -0.07 (-0.2 to 0.07) | 1.90 |
| Discontinuation of maintenance therapy without washout (yes) | -0.08 (-0.1 to -0.03) | 0.3 (0.2 to 0.4) | 1.26 |
| Delayed effect (yes) | -0.0005 (-0.06 to 0.06) | 0.3 (-0.4 to 0.9) | 1.92 |

**Table 3.3:** Model selection: Model coefficients for the best model with respect to LOO MSE for each number of covariates. Note that the same models’ heterogeneity and leave-one-out mean squared error are already represented in the previous Figure.

| Nb<br>charact. | EMULATION DIFFERENCE CHARACTERISTICS |  |  |  |  |  |  |  |  |  |  |
| --- | --- | --- | --- | --- | --- | --- | --- | --- | --- | --- | --- |
|  | Intercept | Comparator<br>emulation<br>(good) | Outcome<br>emulation<br>(good) | Mean age<br>difference<br>(centered) | Difference<br>in % female<br>(centered) | Run-in<br>(yes) | Placebo<br>control<br>(yes) | In hospital<br>start (yes) | Dose<br>titration<br>(yes) | Discontin-<br>uation<br>(yes) | Delayed<br>effect<br>(yes) |
| 0 | 0.002 | - | - | - | - | - | - | - | - | - | - |
| 1 | -0.079 | - | - | - | - | - | - | - | - | 0.282 | - |
| 2 | -0.084 | - | - | - | - | - | - | - | - | 0.287 | 0.338 |
| 3 | -0.115 | - | - | - | - | - | - | 0.203 | - | 0.318 | 0.369 |
| 4 | -0.11 | - | - | -0.011 | - | - | - | 0.245 | - | 0.309 | 0.38 |
| 5 | -0.087 | - | - | -0.01 | 0.005 | - | - | 0.152 | - | 0.357 | 0.318 |
| 6 | -0.098 | - | - | -0.011 | 0.006 | - | - | 0.116 | 0.042 | 0.375 | 0.326 |
| 7 | -0.119 | - | - | -0.011 | 0.006 | - | 0.024 | 0.127 | 0.054 | 0.395 | 0.324 |
| 8 | -0.16 | - | 0.051 | -0.011 | 0.006 | - | 0.03 | 0.127 | 0.046 | 0.402 | 0.331 |
| 9 | -0.112 | -0.025 | 0.029 | -0.011 | 0.008 | 0.127 | - | 0.089 | 0.06 | 0.308 | 0.313 |
| 10 | -0.1 | -0.037 | 0.029 | -0.011 | 0.008 | 0.138 | -0.012 | 0.089 | 0.059 | 0.296 | 0.313 |

The best model with three emulation difference characteristics includes delayed effect, discontinuation and in hospital start. This model’s residual heterogeneity is 1.003. Figure 3.5.B shows the association between the combination of these finally selected characteristics and the outcome - the difference in log hazard ratio. Only the prediction intervals for the combinations with observations are displayed, *e*.*g*., none of the trials in RCT-DUPLICATE has more than one of the three emulation differences set to “Yes”. The three included characteristics are mutually exclusive and together they were better in reducing observed heterogeneity than “close emulation”. Hence, the remaining characteristics only added noise to the indicator for “close emulation”, or they canceled each other out.

## 4 DISCUSSION

Using data from the RCT-DUPLICATE initiative comparing results from RCT-RWE study pairs we observed that the study emulation characteristics “delayed effect of treatment”, “discontinuation during run-in period” and “in-hospital start of treatment”, explained most of the observed variation beyond chance in this sample. In this collection of RCT-RWE study pairs, most of the observed variation in effect estimate differences could be explained by those three emulation characteristics. Surprisingly little variation is explained by “placebo comparator” which was thought to be an emulation challenge in the absence of placebo in clinical practice and source for confounding bias.

It has to be noted that even though RCTs are seen as the standard in establishing the efficacy of medical products they may neither be free of flaws in their implementation, nor might they always represent clinical practice. The results of multiple clinical trials that address similar questions, even basically identical “twin trials” can vary in their findings (see for example [35] and [36], or [37] and [38], or [39]). Discordant results between RCTs and RWE studies that study similar drug exposures and outcomes should not necessarily discredit the RWE study before considering emulation differences that may result in studying a slightly different causal question. Therefore, emphasis has to be put on understanding where those differences come from, and the clinical or research question that is being asked by each study type.

Our study has some limitations. First, we present the results of an exploratory analysis using a limited sample size from 29 RCT-RWE study pairs non-randomly selected by in the RCT-DUPLICATE initiative. Therefore, we could only include a limited number of explanatory emulation characteristics in our models. Second, the trials that were included in RCT DUPLICATE were selected to have high potential to be feasible to emulate with claims data. Relatedly, the design emulation challenges that were recorded may not be a comprehensive listing of all important design differences that could be considered. Different design emulation differences might be more or less relevant for different clinical areas and the direction of the effect of these differences are context dependent limiting the generalizability of our empirical findings.

Overall, our study demonstrates that a substantial proportion of the observed variation between results from RCTs and RWE studies can be attributed to design emulation differences. Furthermore, our study shows how meta-regression can be used to get a more nuanced understanding regarding emulation differences.

## Data Availability

All data and code produced are available online from https://gitlab.com/heyardr/hte-in-rwe.

## Data and code availability statement

Code and data to reproduce the analyses and recompile this manuscript are available through https://gitlab.com/heyardr/hte-in-rwe.

## Ethics approval

The presented study used previously secondary data and therefore no ethics approval was required.

## Transparency statement

The lead author (Dr. Heyard) affirms that this manuscript is an honest, accurate, and transparent account of the study being reported; that no important aspects of the study have been omitted; and that any discrepancies from the study as planned (and, if relevant, registered) have been explained.

## APPENDIX Details on statistical analyses

Our statistical analyses require that the effect estimates are approximately normally distributed around the true effect size *θ*. A log-transformation is applied on the hazard ratios resulting in 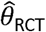 being the estimate of the log-hazard ratio of the RCT and 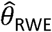 being the (pooled) estimate of the log-hazard ratio of the RWE emulation. The associated variances are denoted by 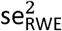 and 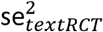 respectively. The standardized differences of each RCT-RWE study pair is computed as such

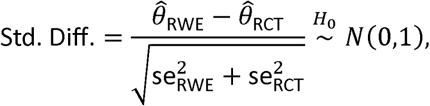

where *H*_0_ is the null hypothesis of absence of heterogeneity and bias. The squared standard difference is the *Q*-statistic with 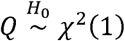 which is used to perform the *Q*-test for heterogeneity between RCT and RWE, often used in meta-analysis [25,26]. Then, 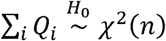 is used for an overall test for heterogeneity between all RCT-RWE study pairs.

The heterogeneity *φ* can be quantified as a multiplicative parameter [27], an overdispersion parameter generally larger than 1 to be applied to each of the trial-specific variances

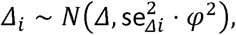

where the variance of the difference of the *i*th study pair is 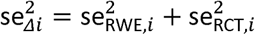. As described in [28], *φ* can be estimated by fitting a weighted linear regression on the observed differences *Δ* _*i*_ from all RCT-RWE study pairs against a constant, with weights 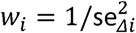. The multiplicative heterogeneity is the model’s standard error truncated at 1 (*φ* = max(*σ*_model_, 1)) and absence of heterogeneity is achieved if *φ* = 1. To explain part of the observed heterogeneity covariates describing emulation challenges (summarised in Table 1) can be used. As in a meta-regression ([29], Chapter 7), the covariates are added into the weighted linear regression model. Then, if the *φ* extracted from the more complex, adjusted model is smaller than the heterogeneity measured with the simple model, we can conclude that part of the variation was explained by the covariates.

### Variable selection

As only a limited number of RCT-RWE study pairs are available, the meta-regression model should be parsimonious to prevent overfitting. According to the commonly used rule of thumb [40] we would need at least ten studies (*i*.*e*., study pairs) for each covariate. To reduce the model complexity, we apply the following variable selection algorithm: all 2^*p*^ possible candidate models are fitted, depending on which combination of the *p* covariates is included. To assess the prediction performance of the candidate models, we compute the leave-one-out cross-validated mean squared error (LOO CV MSE, [30]). This analysis allows us to compare the importance of the covariates and ensure that we select those that are most predictive for our outcome, *Δ* _*i*_ [41]. MSE is oriented in a way that the lower it is, the better the model’s prediction performance. To take the uncertainty of the modeling process into account when selecting a final model, differences in models’ performances are investigated with the interval MSE ±1 · SE, where SE is the LOO standard error. The final selected model will be the smallest model (*i*.*e*., the model with the fewest covariates) with MSE ≤ 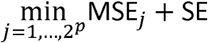 [31].

